# Adherence in Monitoring of ART response and turnaround time of results as per HIV viral load testing guideline among people living with HIV in Dar es salaam Region

**DOI:** 10.64898/2026.04.14.26350908

**Authors:** Tumaini Masegese, Gasper Singfrid Mung’ong’o, Benjamin Kamala, Amani Anaeli, Mussa Bago, Mtoro J. Mtoro

## Abstract

**Background:** HIV/AIDS remains a major public health challenge in Tanzania, where viral load suppression among adults on ART stands at 78% and HVL testing uptake among eligible patients is approximately 22%. Since the introduction of the National HVL Testing Guideline in 2015, little has been done to systematically evaluate its implementation.

**Objective:** To evaluate adherence to the National HVL Testing Guideline across CTC clinics in Dar es Salaam Region, covering ART monitoring, documentation, turnaround time, and factors affecting implementation.

**Methods:** A cross-sectional study was conducted in 2021 across 15 public health facilities with CTC clinics in all five Dar es Salaam districts. A total of 330 PLHIV on ART for more than six months were selected through systematic random sampling with proportional to size allocation, and 45 healthcare providers through convenient sampling. Data were collected via abstraction forms and self-administered questionnaires, and analysed using SPSS Version 23 with descriptive statistics, bivariate analysis, and binary logistic regression.

**Results:** Only 25.1% of patients had their first HVL sample taken at six months as per guideline, with 68.8% delayed beyond six months. Second and third samples were similarly delayed. MoHCDGEC sample tracking forms were absent in 96.7% of facilities and incomplete in 99.1%, and no facility captured specimen acceptance or rejection as site feedback. Turnaround time exceeded the 14-day guideline threshold in 64.5%, 66.7%, and 69.4% of first, second, and third results respectively. Patient negligence (AOR=9.84; 95% CI: 1.83-52.77) and storage (AOR=5.72; 95% CI: 0.94-35.0) were independently associated with guideline adherence.

**Conclusion:** Adherence to the National HVL Testing Guideline in Dar es Salaam is suboptimal across testing timelines, documentation, and turnaround time, with patient negligence and storage capacity as significant determinants. Targeted interventions are needed to strengthen patient education, improve storage infrastructure, enhance documentation systems, and support providers in adhering to guideline-specified timelines.

## Background

The HIV/AIDS pandemic remains one of the most formidable public health challenges worldwide. As of 2024, approximately 40.8 million people were living with HIV (PLHIV) globally, with an estimated 1.3 million new infections and 630,000 AIDS-related deaths during the year (1).

Of all PLHIV, approximately 86% were aware of their HIV status, leaving a gap of roughly 5.4 million undiagnosed individuals worldwide. Progress toward the UNAIDS 95–95–95 targets has reached 87–89–94 globally, with approximately 31.6 million people on antiretroviral therapy (ART) and 73% of all PLHIV having achieved viral load suppression (1).

Sub-Saharan Africa (SSA) continues to bear a disproportionate share of the global HIV burden, accounting for approximately two-thirds of all PLHIV and roughly 60% of new infections worldwide (1). East and Southern Africa is the most severely affected sub-region. Africa’s ten highest-burden countries which include South Africa, Nigeria, Mozambique, Uganda, Tanzania, Zambia, Zimbabwe, Kenya, Malawi, and Ethiopia were collectively responsible for 80% of HIV cases on the continent (2). While significant reductions in new infections and AIDS-related mortality have been achieved through ART scale-up and prevention programming, structural barriers including poverty, gender inequality, stigma, and fragile health systems continue to impede full control of the pandemic (3).

Tanzania continues to face a substantial generalized burden of HIV. According to the Tanzania HIV Impact Survey (THIS) 2022–2023, the prevalence among adults aged 15 years and above stands at 4.4%, translating to approximately 1.55 million people living with HIV. Viral load suppression (VLS) among adults on ART stands at 78%, with notable disparities by sex where suppression is lower among men (72.2%) than women (80.9%), highlighting persistent inequities in treatment outcomes and engagement with HIV care services (4). PLHIV who achieve viral suppression are less likely to develop AIDS-defining conditions and do not transmit HIV to sexual partners, underlining the critical clinical and public health importance of sustained VLS (5,6).

Viral load testing has become the gold-standard approach for monitoring ART effectiveness, supplanting earlier reliance on CD4 cell counts and clinical staging. While CD4 counts provide important information on immunological status, they are poor predictors of virologic failure and cannot detect treatment failure early enough to prevent the accumulation of drug resistance mutations. The World Health Organization (WHO) currently defines virologic failure as a viral load above 1,000 copies/mL on two consecutive measurements after at least six months on ART, with an adherence intervention between measurements (7). Early identification of confirmed virologic failure is paramount to preventing drug resistance accumulation; however, the scale-up of HIV-RNA monitoring in Africa and the timely switch to second-line ART regimens remain challenging (8).

To accelerate progress toward the pandemic control, the WHO introduced a Differentiated Service Delivery (DSD) framework in 2016, with two core goals: providing client-centered services tailored to the needs of different PLHIV populations, and cushioning the impact of increased service demand on overburdened health systems in low- and middle-income countries (9,10). The Universal Test and Treat (UTT) strategy which recommends ART initiation for all PLHIV regardless of CD4 count or clinical stage has been adopted globally and in Tanzania as both a clinical and public health intervention.

A critical component of the DSD framework requires that healthcare providers promptly review and share viral load results with clients, and that clients are educated on the importance of viral load monitoring and viral suppression for their own health and for preventing onward transmission (7).

Tanzania’s national HIV viral load testing guidelines, developed and aligned with WHO recommendations, specify a structured monitoring schedule for all PLHIV on ART. Routine viral load testing is recommended at six months following ART initiation and at 12 months thereafter. For patients whose two preceding results are below 1,000 copies/mL, annual testing is subsequently recommended. For patients with an unsuppressed viral load (≥1,000 copies/mL), the protocol mandates enhanced adherence counselling (EAC) followed by a confirmatory repeat viral load test within three months before any regimen switch is considered (7,11).

A critical operational requirement of the guideline is a turnaround time (TAT) for viral load results of no more than 14 days, with proper documentation of all procedures and results as per guideline standards. To support implementation, Tanzania scaled up its viral load testing laboratory capacity from six to 17 laboratories, serving 1,490 health facilities through a hub-and-spoke specimen referral system (11).

Despite the recognized importance of routine viral load monitoring, uptake and coverage remain suboptimal across much of SSA. A systematic review of the viral load monitoring cascade in SSA documented substantial gaps between recommended and actual testing coverage, with the monitoring cascade encompassing eligibility assessment, specimen collection, laboratory processing, result return, and clinical action presenting multiple points at which patients may be lost (12).

In Tanzania, overall HIV viral load testing coverage has been reported at approximately 22% of eligible ART patients (12). Although viral load testing capacity has expanded considerably, gaps between recommended testing schedules and actual coverage persist, and the implementation of routine viral load monitoring into clinical care has been slower than anticipated (12) .

The gap between guideline recommendation and clinical practice represents a critical implementation challenge. Adherence to national and WHO guidelines for viral load monitoring varies substantially across facilities, regions, and patient populations. Since the introduction of the HVL testing guideline in Tanzania in 2015, little has been done to systematically evaluate its implementation by healthcare providers. Despite the centrality of viral load monitoring to the 95–95–95 targets and to HIV pandemic control, there remains a paucity of studies rigorously assessing real-world implementation of viral load testing guidelines, particularly in high-burden urban settings such as Dar es Salaam. Most existing research has focused on viral load coverage rates or suppression outcomes, with fewer studies examining process indicators of guideline implementation including adherence to recommended testing schedules, turnaround time compliance, documentation practices, and clinical actions triggered by viral load results. The care cascade is a multiplicative model in which gaps at each step compound downstream losses; counting patients with missing viral load data as treatment failures is a key methodological principle to avoid biased and overly optimistic estimates (6).

This study therefore aimed to evaluate adherence to the national HVL testing guideline in monitoring ART response and turnaround time of results among PLHIV in the Dar es Salaam Region, to generate evidence informing targeted quality improvement interventions and to accelerate Tanzania’s progress toward the 95–95–95 targets.

## Materials and methods

### Study design

A cross-sectional study design was conducted among PLHIV on ART treatment more than six months and public health care providers within Care and Treatment Center (CTC) clinics in Dar es Salaam Region. The study retrieved secondary data from the CTC 2 database, which was used to record the clinical management of PLHIV attending CTC. This study design measured exposures and outcome at the same time and hence appropriate for the study.

### Study setting

This study was conducted in Dar es Salaam Region, Tanzania, encompassing all five districts: Kinondoni, Temeke, Ilala, Ubungo, and Kigamboni. The study setting was restricted to public health facilities operating CTC clinics, which provide HIV testing, care, treatment, and prevention services across the region. A total of 116 such facilities were identified, comprising 9 public hospitals, 22 public health centres, and 75 public dispensaries. (13).

Dar es Salaam recorded the highest proportion of discriminatory attitudes toward people with HIV (17.1%), followed by Njombe and Mbeya (both 15%) (14). This elevated discrimination rate may adversely affect HIV viral load testing guideline implementation, making Dar es Salaam the most appropriate study site and underscoring the necessity of this evaluation.

### Study population

The study enrolled male and female PLHIV on ART for more than six months, as this meets eligibility criteria for HVL testing per the National HVL Testing Guideline. Healthcare providers including clinicians, CTC nurses, and laboratory technologists directly involved in HVL testing guideline implementation were also included. All participants were recruited from public health facilities with CTC clinics across each Dar es Salaam district.

### Sampling Procedure

Sample size for PLHIV was calculated using the Kish Leslie formula (1965) for cross-sectional studies, requiring a baseline proportion of PLHIV on ART who had undergone HVL testing. This proportion (22%) was drawn from a Tanzanian study by Antelman (12). For healthcare providers, no sample size calculation was performed; given the small and well-defined population, all providers within CTC clinics at selected facilities were included.

### Sample size calculation

The formula for calculating sample size

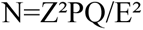

Whereby

N=sample size

Z = standard normal deviation at 95% confidence interval corresponding to 1.96

P = 22% of PLHIV patient eligible on ART who tested for HVL testing (Antelman G et al, 2017)

Q = 1-p

E = margin of error, assumed to be 5%

Therefore

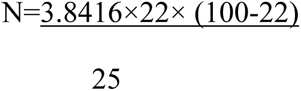

N=264

Assuming the non-response rate = 20%, the minimum sample size, N is **330 respondents.**

### Selection of Facilities

Multistage sampling was used to select 15 public health facilities with CTC clinics. Following the selection of all five Dar es Salaam districts, facilities were selected from each level within each district. Health centres and dispensaries were selected using the lottery method, whereby names of all eligible facilities in each district were written on slips of paper, mixed thoroughly, and drawn without looking. This ensured equal probability of selection for each facility. All district public hospitals were included in their entirety, as each district had only one public hospital, resulting in a final sample of 15 facilities.

### Selection of Participants

Proportional to size sampling was used to determine the number of participants from each CTC clinic. The total number of HIV patients across all 15 sampled facilities was summed, then divided by the sample size (330) to obtain a sampling fraction. This fraction was multiplied by the patient total of each facility to yield the proportional number of participants per clinic.

PLHIV on ART for more than six months were selected through systematic random sampling, with a randomly chosen starting point and every fifth patient identity number selected thereafter. This was applied within each CTC clinic, yielding a total sample of 330, approximately 20 PLHIV per facility.

Healthcare providers including clinicians, CTC nurses, and laboratory technologists were selected through convenient sampling from within the CTC clinics of all 15 sampled facilities.

### Inclusion Criteria

- PLHIV on ART for more than six months
- Clinicians, CTC nurses, and laboratory technologists working at health facilities with CTC clinics

### Exclusion Criteria

- PLHIV with incomplete data in their files

### Study Variables

The dependent variable was adherence to HVL testing guideline implementation. Independent variables included sociodemographic characteristics of healthcare providers, human resources, equipment and supplies, testing space, training on HVL testing, patient negligence, and storage.

### Data collection Method

Data collection was conducted between June and August 2021. Upon visiting each facility, the researcher was introduced to the CTC clinic in-charge and all clinic staff, after which healthcare providers were briefed on the study and invited to participate. Data were collected quantitatively through systematic random review of CTC patient files, covering HVL testing guideline implementation for ART monitoring, documentation of HVL procedures and results, and turnaround time adherence. Findings were recorded on abstraction forms. To examine factors affecting adherence to HVL testing guideline implementation, data were collected via self-administered questionnaires administered to clinicians, CTC nurses, and laboratory technologists. The data collection approach for each variable is summarised in Table 1

**Table 1:**
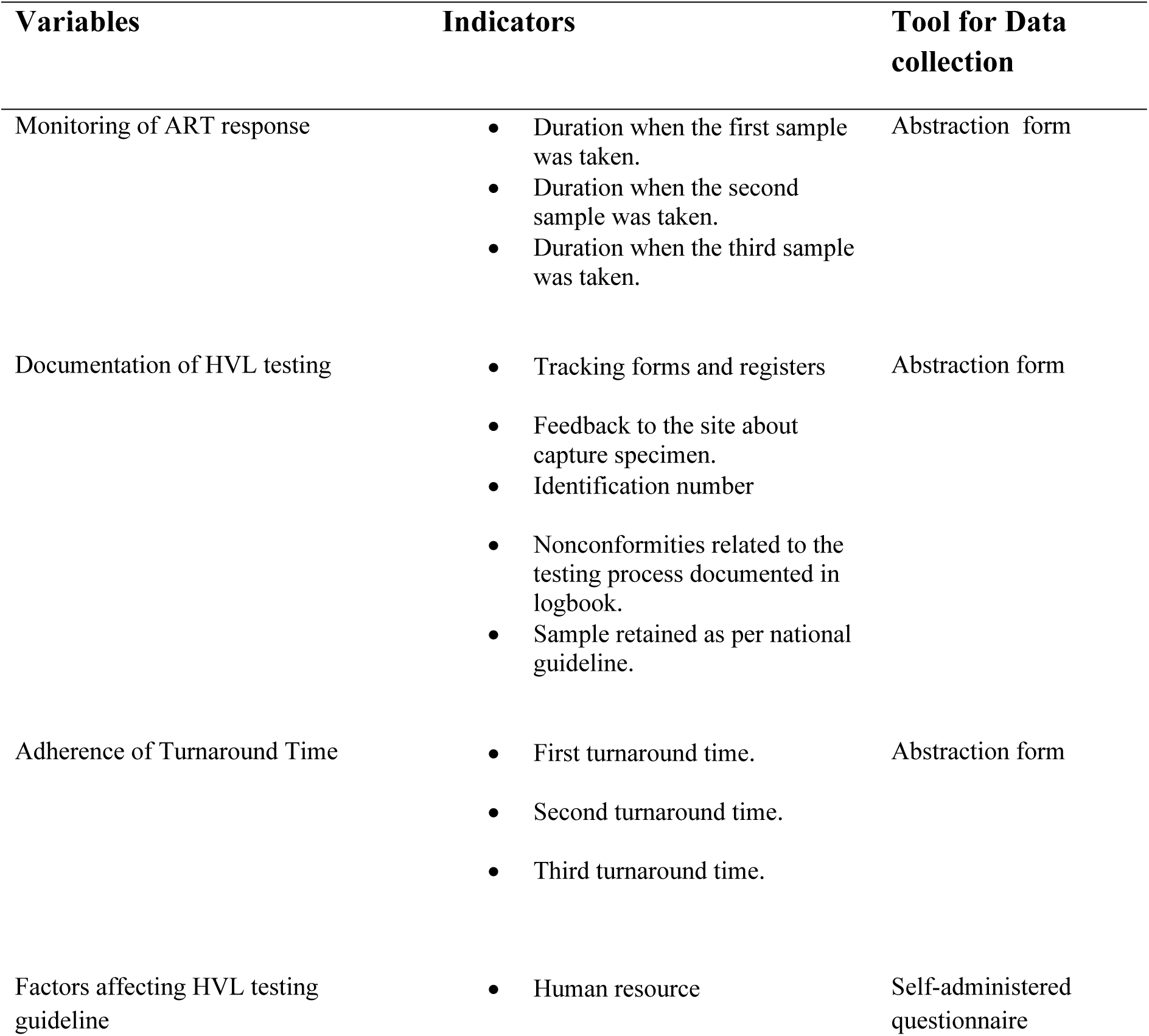
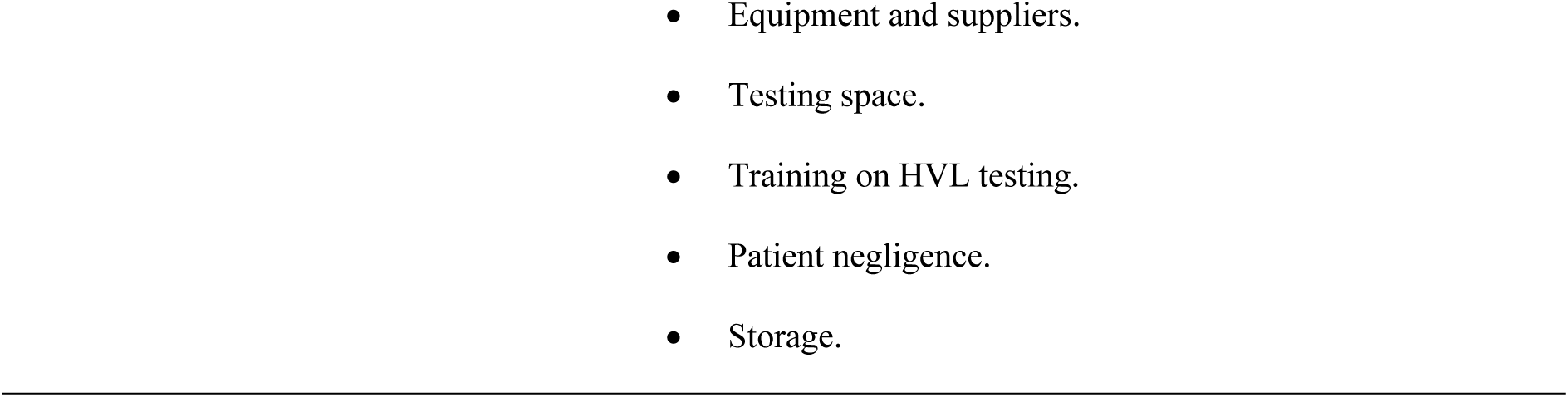
Description on how data were collected based on each variable.

| Variables | Indicators | Tool for Data collection |
| --- | --- | --- |
| Monitoring of ART response | Duration when the first sample was taken.<br>Duration when the second sample was taken.<br>Duration when the third sample was taken. | Abstraction form |
| Documentation of HVL testing | Tracking forms and registers<br>Feedback to the site about capture specimen.<br>Identification number<br>Nonconformities related to the testing process documented in logbook.<br>Sample retained as per national guideline. | Abstraction form |
| Adherence of Turnaround Time | First turnaround time.<br>Second turnaround time.<br>Third turnaround time. | Abstraction form |
| Factors affecting HVL testing guideline | Human resource | Self-administered questionnaire |

### Data Collection Tools

Two tools were used: a researcher-designed data abstraction form for extracting information from CTC patient files of PLHIV on ART for more than six months, and a questionnaire. Both were administered across public health facilities in all Dar es Salaam districts.

### Pre-testing of Tools

A pilot study was conducted at one public health facility in Ilala district to pre-test both tools prior to main data collection. This identified weaknesses in the abstraction form and data collection techniques, and the lessons learned were used to refine the approach.

### Data Analysis Plan

Collected data were checked daily for completeness and correctness, entered into Excel, cleaned, and analyzed using SPSS Version 23 (IBM Corp., Armonk, NY, USA). Both descriptive and inferential analyses were employed.

Frequencies and percentages were used to assess HVL testing across three categories: timing of the first, second, and third samples. Frequencies and percentages described documentation of HVL testing procedures and results. Turnaround time adherence was assessed descriptively across three corresponding time points, determining whether results were returned within 14 days. Descriptive analysis was used to examine factors affecting implementation adherence, followed by bivariate analysis using chi-square to assess associations between independent variables and the dependent variable, with a p-value cut off of 0.05 for inclusion in the regression model. Binary logistic regression was then used to determine statistically significant associations. Education was included in the regression model regardless of p-value to assess its magnitude of association. Findings were presented in narratives, tables, and figures.

### Ethical Considerations

Ethical clearance was obtained from the MUHAS Institutional Review Board (Ref: MUHAS-REC-06-2021-717) which approved data collection for the study. Permission to conduct the study across five Dar es Salaam districts was obtained from each District Medical Officer and District AIDS Coordinator, with further clearance from CTC clinic in-charges. Written informed consent was obtained from all participating clinicians. Confidentiality of all information provided was assured, and data were used solely for the purposes of this study. No personally identifiable information was collected or accessed, and all data were anonymized prior to analysis

## Results

### Socio- demographic characteristics of study participants

A total of 330 patients were studied, out of which 55.8% (n=184) were female. The mean age of was 37.26 years (SD: 10.69), Most of them were between 28-37 years (n=117). Moreover, most were enrolled between 1- 5 years ago 68.2 % (n=225). There was no difference in the district of domicile. Most were found at the dispensary, 139 (42.1%). The details are as shown in Table 1 below.

**Table 1:**
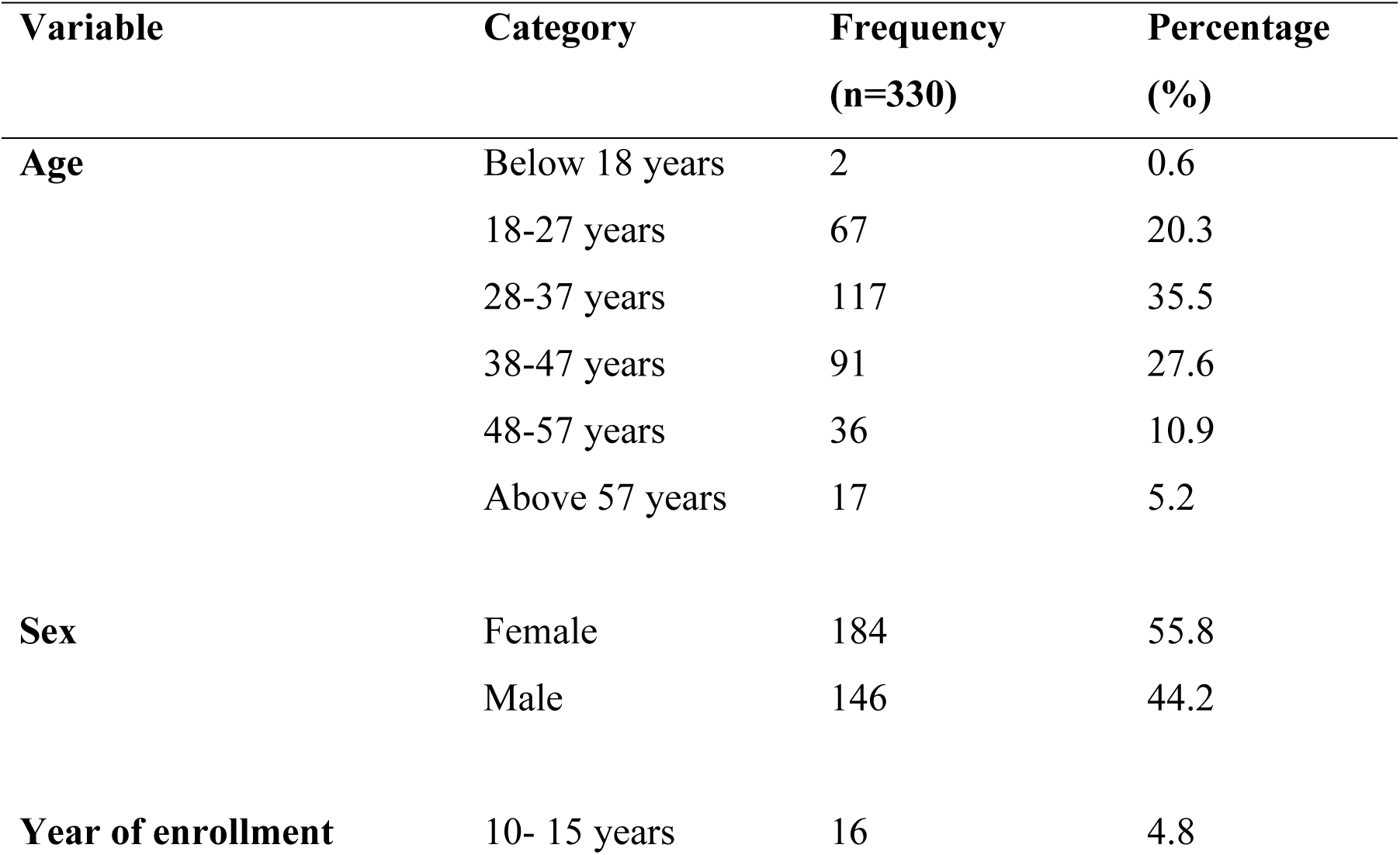
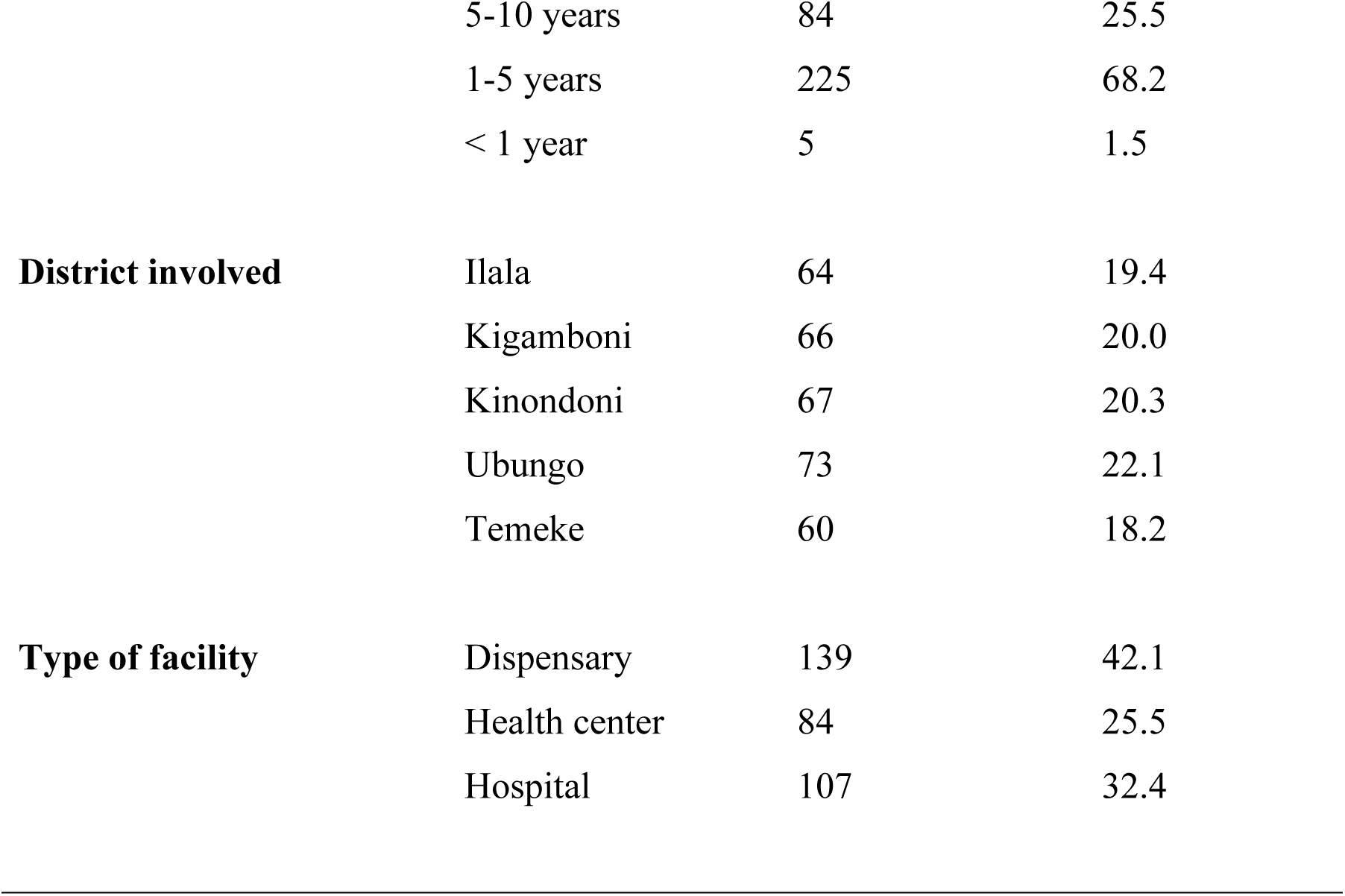
Socio-demographic characteristics of male and female PLHIV on ART (n=330)

### Socio-demographic characteristics of respondent healthcare workers (n=45)

Table 2 illustrates the findings of socio-demographic characteristics of 45 respondents (healthcare providers), whereby more than half were females 31 (68.9%). The mean age of the study participants was 43.27 (SD: 10.50), findings have shown that most of them ranged from 28-37 years, 16 (35.6%) and 38-47 years 16 (35.6%). Additionally, most respondents were from the sampled public hospitals 21 (46.7%), many were nurses 22 (48.9%) and more than half 23 (51.1%) had diploma educational level, with no difference from the districts of domicile.

**Table 2:** Socio-demographic characteristics of respondents (n=45)

| Variable | Category | Frequency<br>(n=45) | Percentage<br>(%) |
| --- | --- | --- | --- |
| Age | 28-37 years | 16 | 35.6 |
|  | 38-47 years | 16 | 35.6 |
|  | 48-57 years | 9 | 20.0 |
|  | Above 57 years | 4 | 8.9 |
| **Cadre** | Nurse | 22 | 48.9 |
|  | Clinician | 19 | 42.2 |
|  | Laboratory technologist | 4 | 8.9 |
| **Sex** | Male | 14 | 31.1 |
|  | Female | 31 | 68.9 |
| **Education** | Certificate | 12 | 26.7 |
|  | Diploma | 23 | 51.1 |
|  | Degree | 10 | 22.2 |
| **District involved** | Ilala | 10 | 22.2 |
|  | Kigamboni | 7 | 15.6 |
|  | Kinondoni | 9 | 20.0 |
|  | Ubungo | 9 | 20.0 |
|  | Temeke | 10 | 22.2 |
| **Type of facility** | Dispensary | 10 | 22.2 |
|  | Health center | 14 | 31.1 |
|  | Hospital | 21 | 46.7 |

### Monitoring of ART response by using HIV viral load testing among CTC clinics

Figure 1 indicates findings based on monitoring of ART response by using viral load testing among CTC clinics of 330 people who are on ART treatment in Dar es Salaam. The findings indicate that 83 (25.1%) of eligible patients were taken first sample at 6 months on ART treatment, 227 (68.8%) eligible patients were taken first sample more than the 6 to 12 months on ART treatment, whereas 20 (6.1%) of eligible patients were taken first sample in more than 12 months.

**Figure 1:**
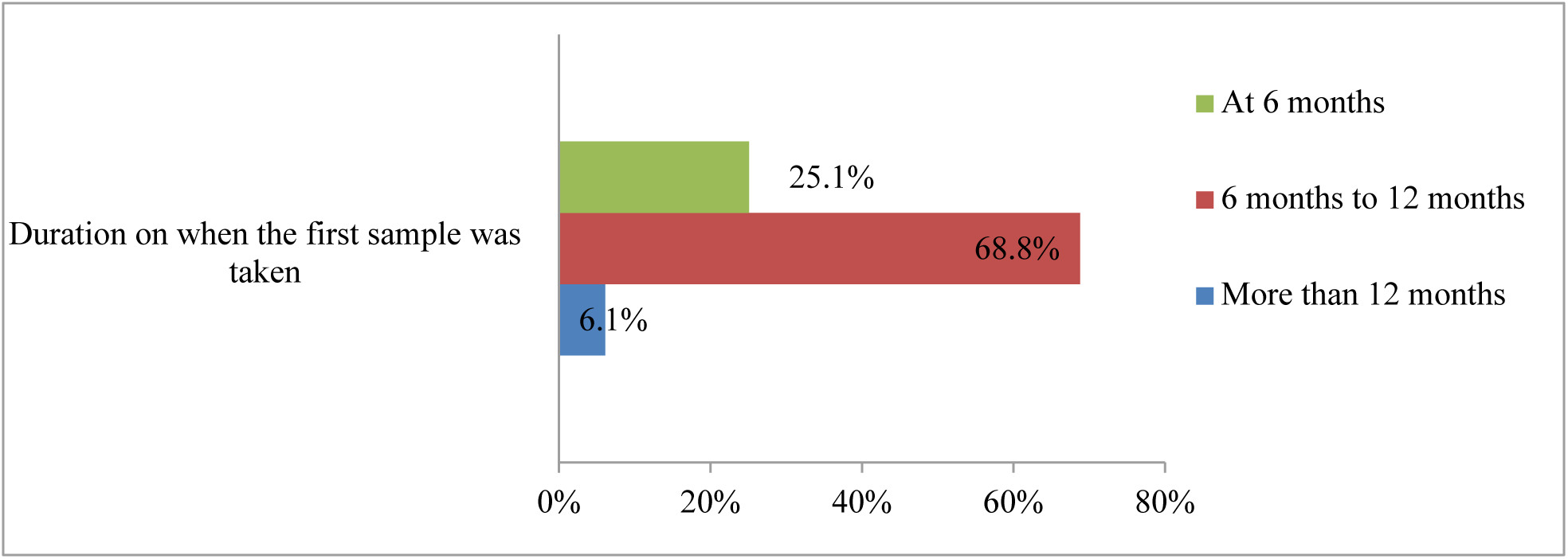
HVL testing among CTC clinics when the first sample was taken (n=330)

Figure 2 also illustrates that among 330 people who are on ART treatment, the majority i.e., 195 (59.1%) had their second HVL sample taken in 12 to 24 months, 116 (35.1) at 12 months whereby the remaining i.e., 19 (5.8%) had their second HVL sample taken more than 24 months.

**Figure 2:**
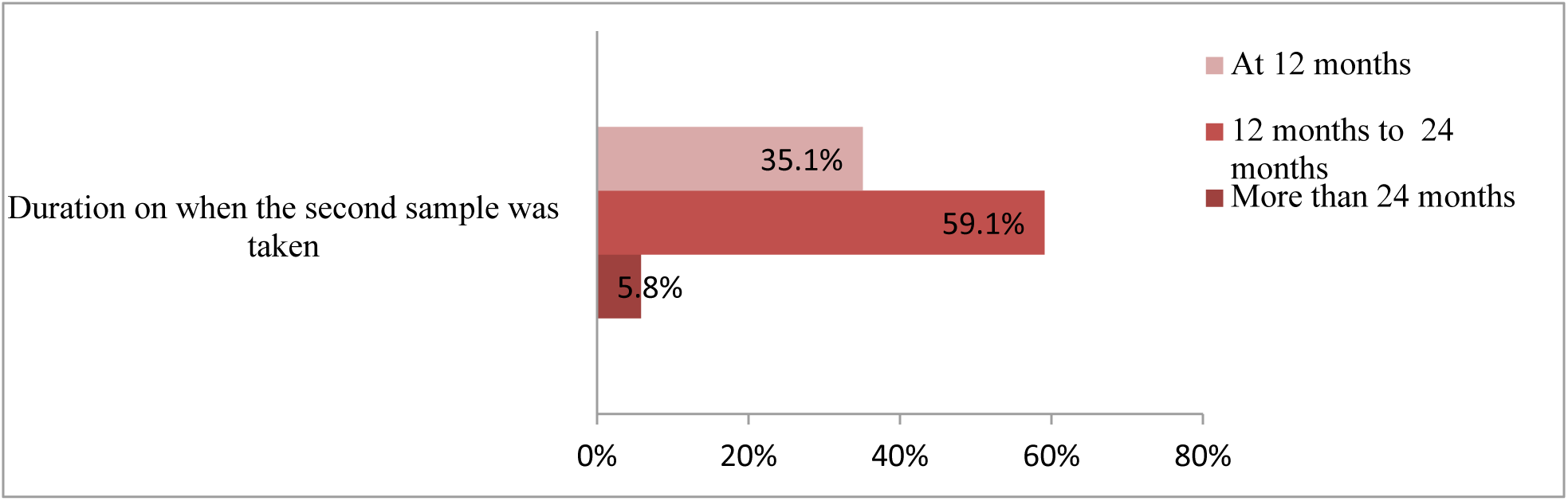
HVL testing among CTC clinics when the second sample was taken (n=330)

The findings in Figure 3 also indicates that among 330 people who are on ART treatment, 224 (67.9%) had their third HVL sample taken in 24 months to 36 months, 60 (18.2%) at 24 months and the remaining 46 (13.9%) had their third sample taken in more than 36 months.

**Figure 3:**
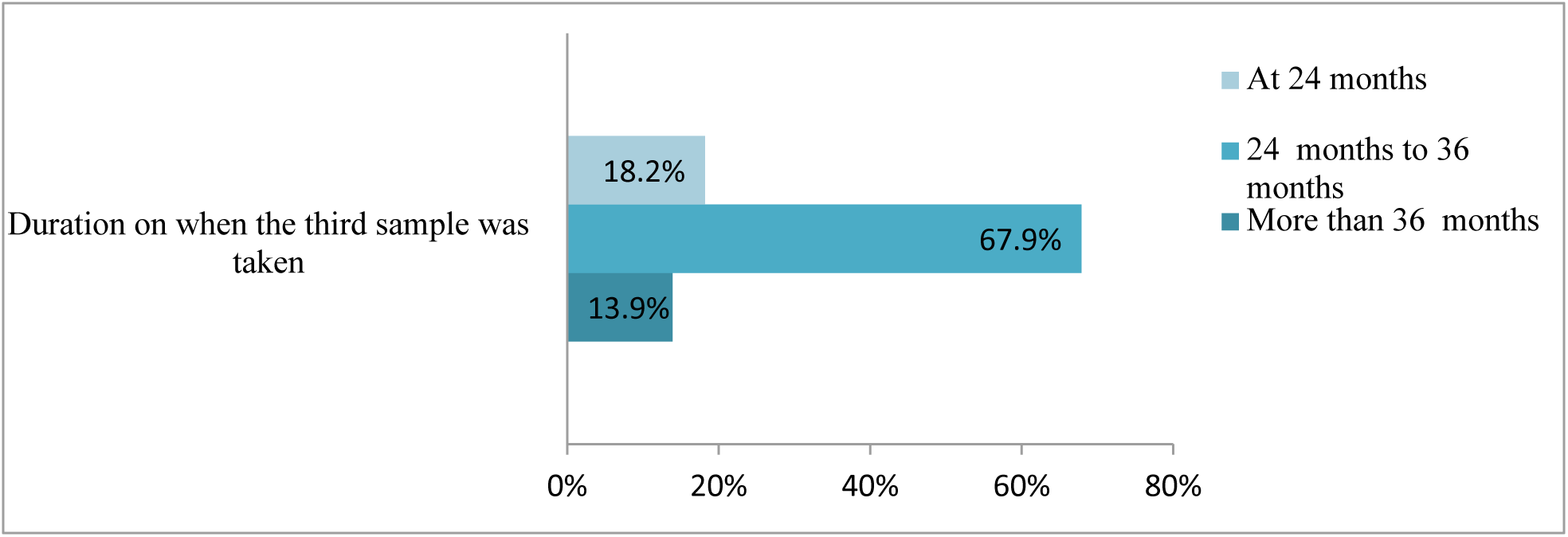
HVL testing among CTC clinics when the third sample was taken (n=330)

### Documentation of HVL testing procedure and results among CTC clinics

Table 3 illustrates the findings based on documentation of HVL testing procedure and results among CTC clinics, the findings indicate the overwhelming majority 319 (96.7%) of standard sample tracking forms by MoHCDGEC were not registered in places, 327 (99.1%) of standard sample tracking forms were found not to be filled completely. The findings based on form capturing specimen acceptance or rejection as feedback to the site show no form capturing specimen acceptance or rejection as feedback to the site of all 330 (100.0%) forms. Moreover, there was not any form 0 (0.0%) used to capture specimens which was not labeled with client identification number. Furthermore, the findings show there was not any form 0 (0.0%) of patients’ test results which was not documented into results register or logbook. Additionally, all 330 (100.0%) forms were found with no nonconformities related to the testing process documented into the logbook. Lastly, the findings in Table 3 illustrate that there was not any 0 (0.0%) samples which was not retained and disposed according to the national guideline.

**Table 3:**
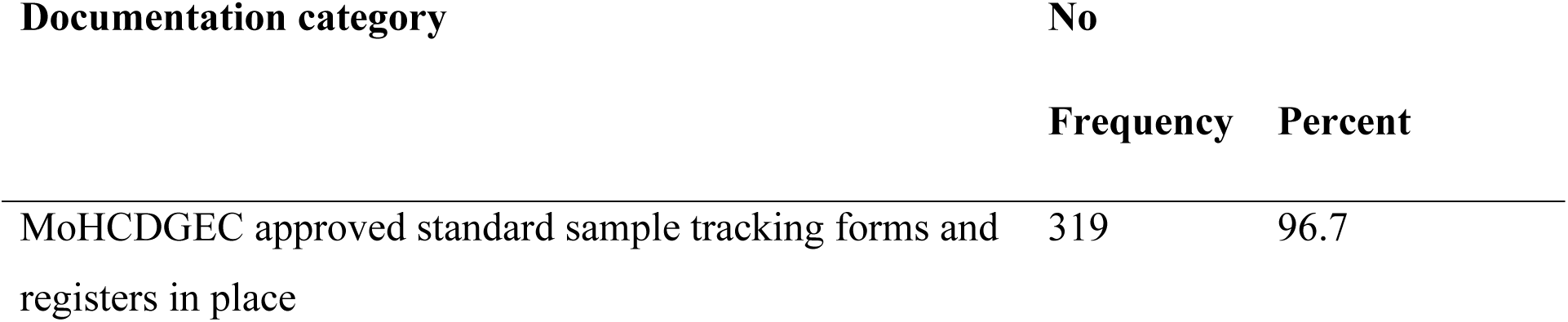
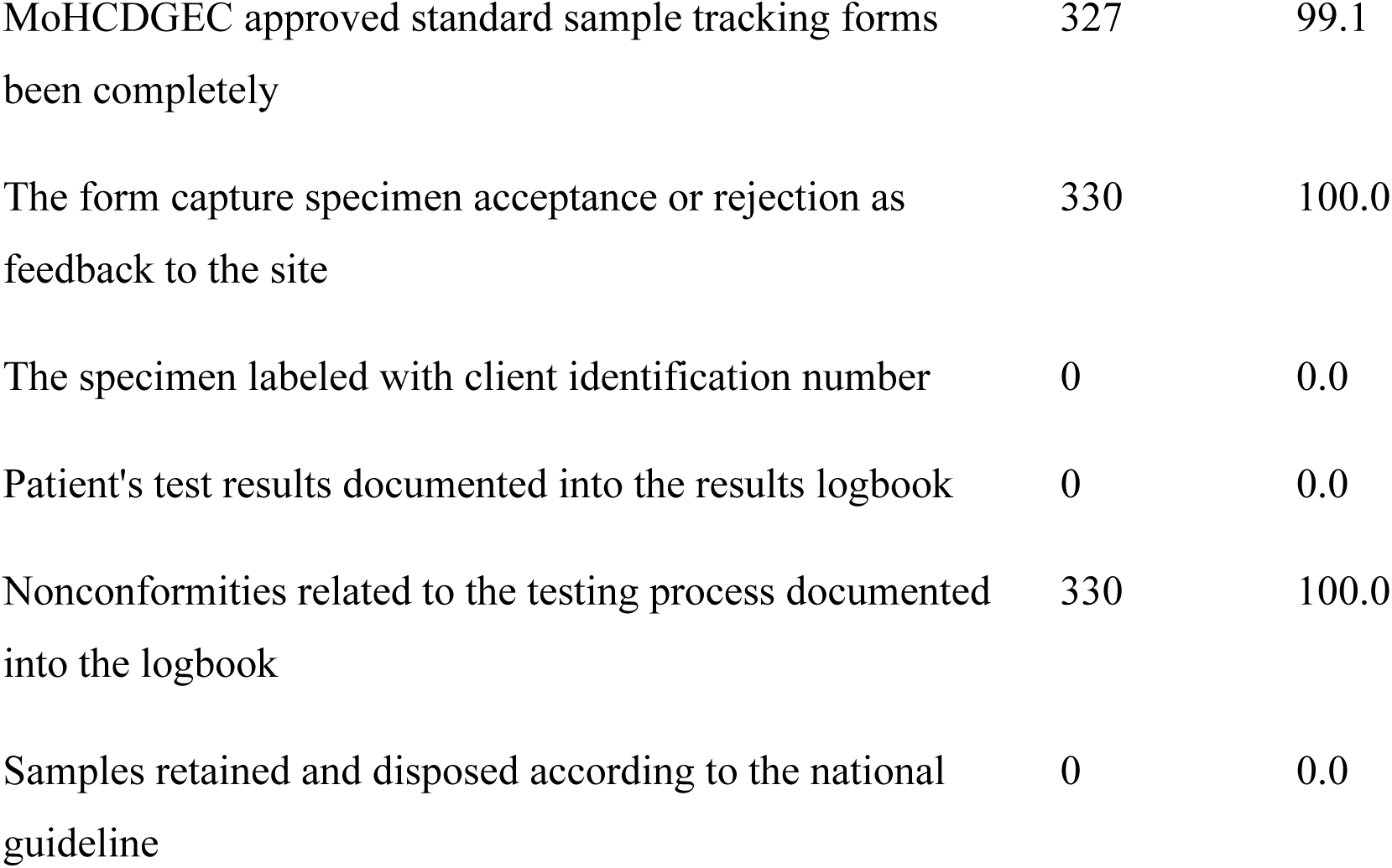
Documentation of HVL testing procedure and results among CTC clinics (n=330)

### The adherence Turnaround of HVL testing results per guideline in CTC clinics

The findings in Figure 4 indicate the adherence Turnaround of HVL testing results per guideline in CTC clinics. Based on a category of first turn around time, majority 213 (64.5%) of reviewed forms showed a turnaround time of delivered with HVL testing results in more than 14 days.

**Figure 4:**
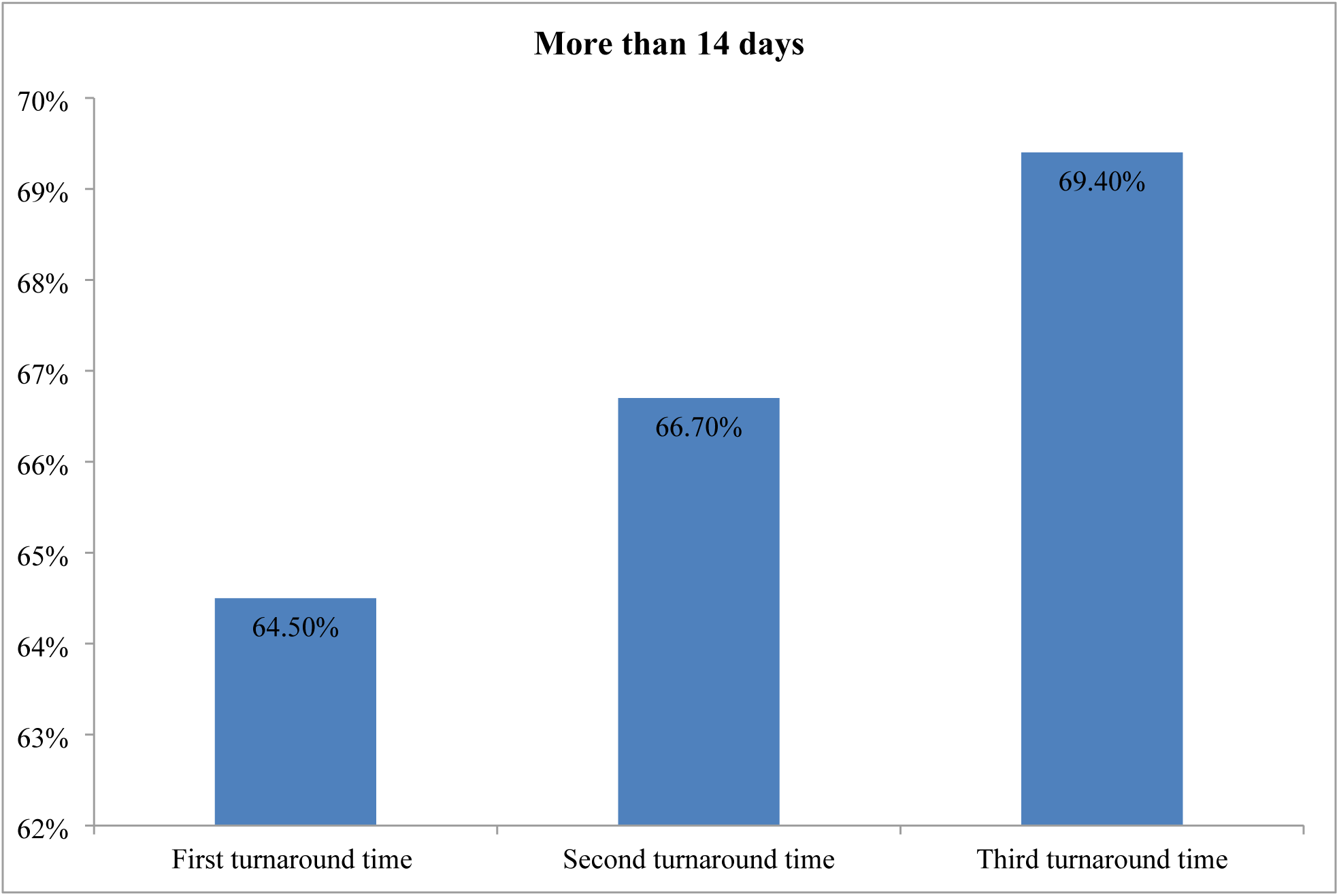
The adherence Turnaround of HVL testing results guideline in CTC clinics (n=330)

Based on a category of second turn around time, the findings in Figure 4 illustrate that 220 (66.7%) of reviewed forms showed a turnaround time of delivered with HVL testing results in more than 14 days.

Lastly, the findings in Figure 4 also indicate that more than half 229 (69.4%) of reviewed form showed to receive a third turn around in more than 14 days.

### Factors affecting implementation adherence to HVL testing guideline

Table 4 presents descriptive findings on factors affecting adherence to HVL testing guideline implementation, based on yes/no responses from 45 healthcare providers. More than half of respondents identified human resource (n=25, 56.6%) and testing space (n=25, 55.6%) as affecting implementation. Equipment and supplies was the most commonly cited factor, with more than three-quarters of respondents (n=34, 75.6%) agreeing it affects implementation. Training on HVL testing and patient negligence were each cited by (n=15, 33.3%) of respondents, while storage capacity was the least cited factor (n=12, 26.7%).

**Table 4:** Factors affecting implementation of HVL testing (n= 45)

|  | **Frequencies** | **Percent** |
| --- | --- | --- |
| **Human resource** (*Human resource is among the factors that affect adherence of HVL testing guideline*) | 25 | 56.6 |
| **Equipment and supplies** (*Implementation of HVL testing guideline is sometimes affected by equipment and supplies*) | 34 | 75.6 |
| **Testing space** (*Testing space affects the implementation of HVL testing guideline*) | 25 | 55.6 |
| **Training on HVL testing** (*In my opinion, training on HVL testing influences implementation of HVL testing guideline*) | 15 | 33.3 |
| **Patience negligence** (*Patience negligence would affect the implementation of HVL testing guideline*) | 15 | 33.3 |
| **Storage** (*Capacity of storage of specimens may affect implementation of HVL testing guideline*) | 12 | 26.7 |

### Factors associated with implementation of HVL testing guideline

During a bivariate analysis patient negligence and storage was significantly associated with implementation of HVL testing guideline (p=<0.001) and (p=0.001) respectively. Health carder showed a marginal association with implementation of HVL testing guideline (p=0.067). Of 7 (50%) of the participant aged 48+ years were implementing HVL testing guideline and the similar pattern was observed in health facility were 5 (50%) of dispensaries were implementing HVL testing guideline as shown in Table 5. 7 (58.33%) of participants having certificate education were implementing HVL testing guideline. Training on HVL testing was not significantly related with implementation of HVL testing guideline (P=0.138).

**Table 5:** Bivariate analysis of factors associated with Implementation of HVL testing guideline (N=45)

| Characteristics | Total<br>N | Implementation of HVL<br>testing guideline n (%) | | $\chi^2$ (df) | P value |
| --- | --- | --- | --- | --- | --- |
|  |  | Yes | No |  |  |
| **Age (in years)** |  |  |  |  |  |
| 28-37 | 16 | 5 (31.25) | 11 (68.75) |  | 0.255* |
| 38-47 | 15 | 3 (20) | 12 (80) |  |  |
| 48+ | 14 | 7 (50) | 7 (50) |  |  |
| **Health Facility** |  |  |  |  |  |
| Dispensary | 10 | 5 (50) | 5 (50) |  | 0.358* |
| Health Center | 14 | 3 (21.43) | 11 (78.57) |  |  |
| Hospital | 21 | 7 (33.33) | 14 (66.67) |  |  |
| **Education** |  |  |  |  |  |
| Certificate | 12 | 7 (58.33) | 5 (41.67) |  | 0.098* |
| Diploma | 23 | 5 (21.74) | 18 (78.26) |  |  |
| Degree | 10 | 3 (33.33) | 7 (70) |  |  |
| **Health Cadre** |  |  |  |  |  |
| Nurse | 22 | 11 (50) | 11 (50) |  | 0.067* |
| Clinician | 19 | 4 (21.05) | 15 (78.95) |  |  |
| Lab tech | 4 | 0 | 4 (100) |  |  |
| **Human resource** |  |  |  |  |  |
| Yes | 25 | 10 (40) | 15 (60) | 1.13 (1) | 0.289 |
| No | 20 | 5 (25) | 15 (75) |  |  |
| **Equipment** |  |  |  |  |  |
| Yes | 34 | 11 (32.35) | 23 (67.65) |  | 1.000* |
| No | 11 | 4 (36.36) | 7 (63.64) |  |  |
| **Testing space** |  |  |  |  |  |
| Yes | 25 | 9 (36) | 16 (64) | 0.18 (1) | 0.671 |
| No | 20 | 6 (30) | 14 (70) |  |  |
| **Patient negligence** |  |  |  |  |  |
| Yes | 15 | 11 (73.33) | 4 (26.67) | 12.6 (1) | <0.001 |
| No | 30 | 4 (13.33) | 26 (86.67) |  |  |
| **Storage** |  |  |  |  |  |
| Yes | 12 | 9 (75) | 3 (25) |  | 0.001* |
| No | 33 | 6 (18.18) | 27 (81.82) |  |  |
| **Training** |  |  |  |  |  |
| Yes | 25 | 6 (24) | 19 (76) | 2.21 (1) | 0.138 |
| No | 20 | 9 (45) | 11 (55) |  |  |

**\*fisher exact p-value**

Patient negligence was significantly associated with implementation of HVL testing, whereas those who adhered to Implementation of HVL guideline were 17.88 times more likely to agree that patient negligence affected HVL testing guideline (OR=17.88; 95% CI: 3.78-84.63). Those who adhered to implementation of HVL guideline were 3.27 times more likely to be certificate holder than degree holder (OR=3.27; 95% CI: 0.55-19.35). Those who adhered to implementation of HVL guideline were 13.5 times more likely to agree that storage affected HVL testing guideline (OR=13.5; 95% CI: 2.79-65.40).

During multivariate analysis, those who adhered to implementation of HVL guideline were 9.84 times more likely to agree that patience negligence affected HVL testing guideline (AOR=9.84; 95% CI: 1.83-52.77). Moreover, those who adhere to Implementation of HVL guideline were 5.72 times more likely to agree that storage affected HVL testing guideline (AOR=5.72; 95% CI: 0.94-35.0).

**Table 6:**
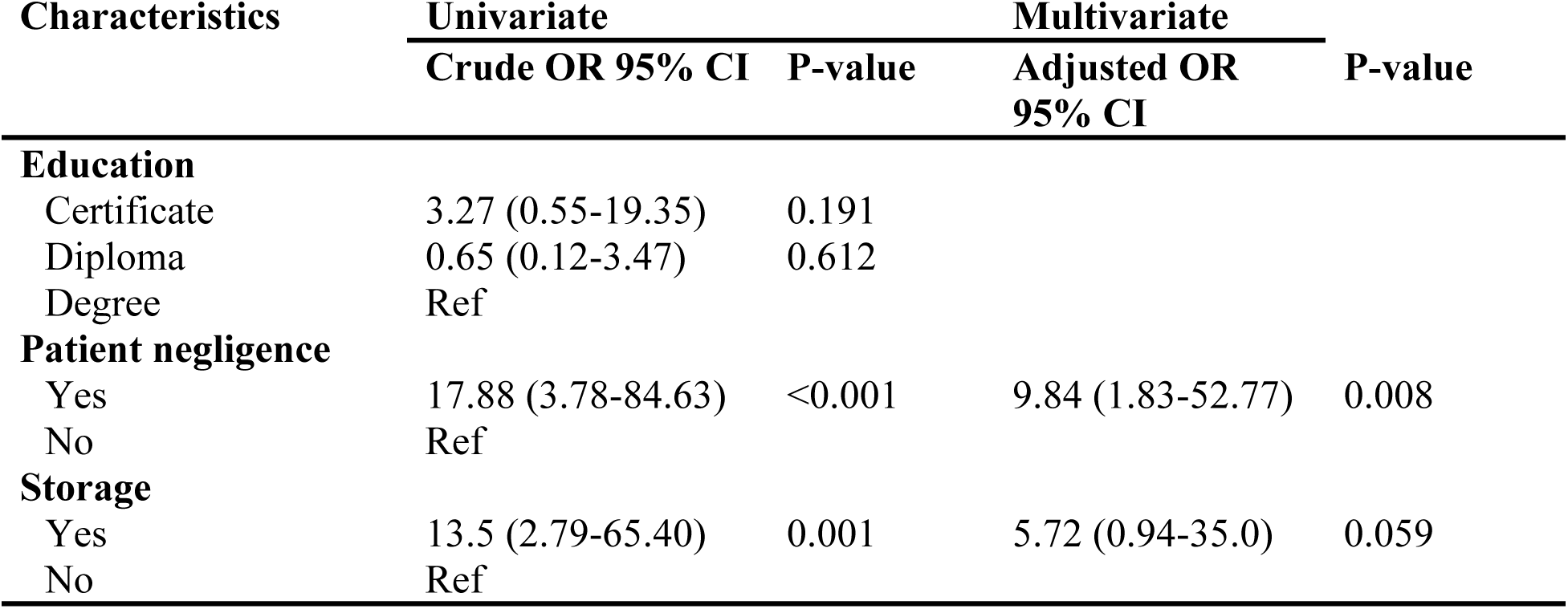
Logistic Regression model of factors associated with implementation of HVL testing guideline.

## Discussion

The findings based on monitoring of ART responses by using HIV load testing among CTC clinics imply that, there is a delay of majority of patients who are in ART treatment to be taken their HVL sample testing. Most of the patients were found not to be taken their first HVL testing according to the guideline time (six months), instead the first sample was taken in more than 6 months. In this study only less-half of the patients had their first HVL sample testing as per guideline. This conveys effect to delay of the second and third HVL sample testing. In this case, most of the patients did not get immunological diagnosis treatment failures due to a delay in taking HVL testing. Furthermore, the findings are comparable with the findings reported in the study done in United Kingdom on HIV transmission rates from persons living with HIV who are aware and unaware of their infection their study found that more than half of HIV patients increase the burden of disease because of not taking HVL testing timely (15). The reported findings are seen to be similar maybe because little of awareness of the patients when they are registered for first time for HVL testing in both areas where the authors have done their studies, for instance, in Tanzania this study has proven that most of the patients are not aware of the consequences of delaying for HVL testing.

Moreover, the situation observed in the duration of when the first sample was taken has been the same from when the second sample was taken, as it was shown that most of the patients who were on ART treatment more than six months their second HVL testing was also delayed not as per guideline (12months). These findings showed that there is still poor adherence on implementation of HIV load testing guideline. However, the findings diverge from the one reported in Tanzania and found that only 33% of people with HIV who are on ART treatment were more likely to be tested on second line of their ART treatment (12). Moreover, the obtained findings also distinct from the results obtained in the study conducted in Myanmar, the results showed that after 6 months of initiation 87.1% of ART retained in care and were eligible for routine HIV load testing. Also among them 57.8% got HIV load testing in six months, additionally 75.0% got HIV load testing at second duration time in one year. This means in both duration (first and second duration when sample was taken) the patients never delayed (16). The findings seem to vary in different studies, because of efforts done in each country independently to create awareness to both healthcare providers and patients, in the countries were more efforts are done the impact is to improve adherence of HVL testing.

The results also showed 18% patients had their third sample was taken which is less half of the population. More than half of patients (67.9%) attended for ART treatment in 24 to 36 months for their third HVL sample. Furthermore, the findings is contrary to the study done in Dodoma which reported and argued that when the third sample is taken, a minimum of eleven months and maximum of twelve months (1 year) should be considered for another ART treatment and HIV load testing (17). Here, the findings are seen to be similar, because all these two studies have been done in Tanzania, where the alike level of efforts to encourage healthcare providers to improve their adherence of HVL testing by following the guideline.

The findings demonstrate ineffective documentation of ART treatment. Among the seven items used to assess documentation in the visited health facilities, only three items were found to be done properly as per HIV load testing guideline. These three items include labeling specimens with clients’ identification number, patients’ test results documented into the results registered or logbook and retaining and disposing samples according to the national guideline. These findings of the three items were reflecting on the HIV load testing guideline. This makes clear that health care providers should make sure all information are accurate and filled correct in the register logbook for tracking implementation of ART treatment. The findings were also similar to the study reported in Tanzania in 2017, which showed patients registered in files and database timely helps to identify number of clients on ART treatment < 6 and see if they had received VL tests as per National guideline (18). The findings are found to be similar because the matter registering is very significant when a patient starts to undergo HVL testing; it is not possible to provide services without tracking records or information from patient’s file.

Apart from the three mentioned items which were found to be considered in documentation of HVL testing, there are other four items which have shown the gap in the issue of documentation. The gaps in these four items include MoHCDGEC approved standard sample tracking forms and registers were not in place, incomplete approved standard sample tracking forms by MoHCDGEC, undelivered feedback of form used to capture specimen acceptance or rejection, and undocumented of nonconformities related to the testing process into logbook. Additionally, these findings are in line with the findings found in study done in Ghana, which states that there has been reported incidence of gaps on the issues of documentation of HVL testing in African region particularly in Ghana. Therefore, his arguments are not far from what has been obtained in this study, the findings have proven that in the study area there is also a gap in documentation, as some of documentation activities are considered as per HVL testing guideline but other activities are not considered(19).

The current study provides information on turnaround time of HVL testing as per guideline in CTC. It is observed in the findings that to a large extent there is no adherence to turnaround time of the HVL results. Similar findings were reported in a study done in South Africa to identify barriers to implementation of point of care testing, among the identified barriers was long test turnaround time (20). This calls for urgent attention to treatment monitoring and especially on adherence to turnaround time among patients on ART more than six months. Moreover, the results are similar because most of developing countries including Tanzania and South Africa were the apprehensive studies have been conducted, there is shorten of healthcare providers, this may lead to the delay of turnaround time of HVL testing. It is important to adhere to turnaround time so as to avoid delays in switching to second-line ART and poor health outcomes including prolonged immune activation, development of drug resistance and risk of opportunistic infections.

This study examined different factors affecting the implementation of adherence to HVL Testing guideline among CTC clinics. The factors included human resource, equipment and supplies, testing space, training on HVL testing, patient negligence and storage. Also, the findings of the study have shown patient negligence and storage were significantly associated with implementation of HVL testing guideline. These findings are similar to the study in Myanmar, which reported on the uptake of VL testing and factors associated with it (16). However, the findings are contrary with the study done in South Africa, which showed the significant factors with the implementation of ART treatment according to the guideline to be marital status, being away from home, forgetfulness, non-discloser and alcohol use (21). Also the difference is, the current study has obtained few significant associated factors due to small study sample size compared to the study done in Myanmar, which had factors such as limited human resources in facilities, lack of training and difficulties in sample transportation (16). This information embolden that ART clinics has to be equipped with storage facilities or access to storage, but also provision of knowledge on HVL testing to patients to reduce patient negligence and improve the implementation of HVL testing guideline.

## Study Limitations

This study had a few limitations worth acknowledging. Due to time constraints, PLHIV on ART were not interviewed; their perspectives on the National HVL Testing Guideline would have enriched the findings, and future studies should therefore incorporate this population. The use of convenient sampling to select healthcare providers is another limitation, as findings from this group cannot be generalised to broader populations beyond the study region or district. Regarding data collectors, recruited enumerators did not initially possess adequate knowledge and skills for data collection; however, this was mitigated through structured training conducted by the researcher prior to fieldwork, equipping all data collectors with the necessary knowledge and interview techniques before proceeding to the field.

## Conclusion

This study concludes that monitoring of ART response through HIV viral load testing across CTC clinics in Dar es Salaam region is not being carried out effectively in accordance with the National HVL Testing Guideline. Patients on ART treatment experience delays in having their HVL samples collected across all three testing points stipulated by the guideline. Only a small proportion had their first sample taken at six months from ART initiation as required. Although adherence to testing timelines shows some improvement with the second and third samples, the overall lack of consistency complicates effective monitoring of ART response and undermines the objectives of the guideline.

With respect to documentation, the study finds that while certain documentation activities are conducted in line with the guideline, a substantial number are not. Specifically, MoHCDGEC sample tracking forms were frequently found to be incomplete, and there was no systematic capturing of specimen acceptance or rejection as feedback to the site, representing a significant gap in the documentation process.

Regarding turnaround time, the study confirms that results are not being delivered within the guideline-stipulated 14 days across all three turnaround time points. The delays observed were consistent and pervasive, affecting the first, second, and third turnaround times alike, which has direct implications for timely clinical decision-making for patients on ART.

Finally, this study establishes that a statistically significant association exists between certain factors and adherence to HVL testing guideline implementation. Of the six factors examined, four were found to directly influence adherence: education, health cadre, storage, and patient negligence. These findings provide an evidence base for targeted interventions to improve guideline adherence in similar settings.

### Recommendations

Given that HVL testing for ART monitoring is not being conducted effectively, concerted efforts should be made to ensure that all patients on ART for more than six months are tested as per the guideline. Healthcare providers should also routinely provide health education to patients on the importance of HVL testing in monitoring ART response, as this may reduce instances of patient negligence during sample collection.

In view of the documentation gaps identified, health system managers in the study area are recommended to develop and implement an enhanced specimen tracking system that supports accurate and timely documentation. Such a system would help identify and address inefficiencies in real time.

To address turnaround time delays, healthcare providers should be supported and held accountable in adhering to guideline-specified timeframes. Facilities should be adequately equipped with all necessary supplies and equipment to enable timely processing and delivery of HVL testing results to patients.

Since the study has demonstrated that specific factors significantly affect adherence to HVL testing guideline implementation, the government and relevant health authorities should invest in targeted solutions including expansion of facility capacity, improvement of specimen storage infrastructure, strengthening of health education for patients, scale-up of HVL testing training for healthcare providers, and procurement of adequate equipment and supplies.

Future studies should consider qualitative approaches to obtain a broader and deeper understanding of the factors affecting HVL testing guideline implementation among healthcare providers. Additionally, PLHIV on ART who are eligible for HVL testing should be included as study participants to capture their experiences and perspectives on the National HVL Testing Guideline.

## Data Availability

Due to ethical and confidentiality restrictions regarding sensitive HIV patient data, the dataset is not publicly available. Data are available upon reasonable request to the corresponding author.

## Acknowledgements

The authors express their sincere gratitude to all who contributed to the success of this study. Special appreciation is extended to Dr. Benjamin Kamala and Dr. Amani Anaeli for their guidance and supervision throughout this work. We are grateful to the District Medical Officers and District AIDS Coordinators of all five districts in Dar es Salaam Region, namely Kinondoni, Temeke, Ilala, Ubungo, and Kigamboni, for granting permission to conduct the study and for their cooperation. We also thank the in-charges and staff of all CTC clinics within the 15 sampled public health facilities, spanning district hospitals, health centres, and dispensaries, for their willingness to support and participate in this research. Sincere appreciation is extended to all data collectors for their dedication during fieldwork. This study was conducted as part of a Master of Science degree in Project Management, Monitoring, and Evaluation in Health at Muhimbili University of Health and Allied Sciences.

## Author Contributions

- Conceptualization: Tumaini Masegese
- Data curation: Tumaini Masegese, Mtoro J. Mtoro
- Formal analysis: Mtoro J. Mtoro
- Funding acquisition: Tumaini Masegese
- Investigation: Tumaini Masegese
- Methodology: Tumaini Masegese, Amani Anaeli, Benjamin Kamala
- Project administration: Tumaini Masegese
- Supervision: Amani Anaeli, Benjamin Kamala
- Validation: Amani Anaeli, Benjamin Kamala, Musa Bago,
- Writing – original draft: Tumaini Masegese, Gasper Singfrid Mung’ong’o
- Writing – review & editing: Amani Anaeli, Benjamin Kamala, Musa Bago

## Notes

### Competing Interest Statement

The authors have declared no competing interest.

### Funding Statement

The authors received no specific funding for this work.

### Author Declarations

Ethical clearance was obtained from the MUHAS Institutional Review Board (Ref: MUHAS-REC-06-2021-717)

